# Measures of Street Drug Availability and US Drug Overdose Mortality in the Era of Fentanyl and Stimulants

**DOI:** 10.1101/2023.08.08.23293837

**Authors:** Manuel Cano, Patricia Timmons, Madeline Hooten, Kaylin Sweeney

## Abstract

**Background:** Street-sold drugs such as illicitly-manufactured fentanyl and stimulants have replaced prescription opioids as the primary contributors to fatal overdoses in the United States (US), yet the street availability of these substances is challenging to quantify. Building on the foundation of prior research on law enforcement drug seizures, the present study compares a variety of publicly-available drug seizure measures to identify which measures account for the most variation in drug overdose mortality between states, within states over time, and in various US demographic groups.

**Methods:** Drug seizure counts from the National Forensic Laboratory Information System and drug overdose mortality rates from the Centers for Disease Control and Prevention were examined for all US states, 2013-2021 (459 state-years). State- and year-fixed effects models regressed drug overdose mortality rates (in the overall population and subpopulations by sex, age, and race/ethnicity) on various drug seizure measures, including rates per population and proportional shares of fentanyl-related, heroin, cocaine, methamphetamine, and xylazine seizures.

**Results:** For drug overdose death rates in the overall population and all subpopulations examined by sex, race/ethnicity, and age (except ages 15-29), the model including all drug seizure proportional measures represented the best-performing model examined (as identified via the lowest Akaike Information Criterion and highest within R-squared value), followed by the model including only the fentanyl-related seizure proportion.

**Conclusions:** Findings support the utility of publicly-available drug seizure composition measures, especially the proportion of fentanyl-related seizures, as potential proxies for street drug availability across the US and in various subpopulations.

## 1. Introduction

Illicitly-manufactured drugs such as non-pharmaceutical fentanyl and stimulants have replaced prescription opioids as the drugs most frequently involved in fatal overdoses in the United States (Spencer et al., 2022). In 2021, for example, synthetic opioids (e.g., fentanyl) were reported in 70,601 overdose deaths, psychostimulants (e.g., methamphetamine) in 32,537 deaths, and cocaine in 24,486 deaths; in contrast, prescription opioid pain relievers were reported in 13,618 deaths (Spencer et al., 2022). The shift from prescription opioids to illicitly-manufactured drugs has required researchers to identify measures to assess the availability of these drugs (Hall et al., 2021; Jalal and Burke, 2021; McBrien and Alexander, 2022; Rosenblum et al., 2020) in order to better understand, and respond to, ever-changing trends in overdose deaths.

Law enforcement drug seizures represent one available source of measures related to the illicit drug supply and fentanyl availability. Several recent studies have examined associations between drug seizures and measures of overdose mortality or have developed methods of using drug seizure data to forecast or predict overdose deaths as part of early-warning systems. Many of these studies have focused on data from a single state or area (e.g., Hall et al., 2021; Mohler et al., 2021; Lowder et al., 2022; Ray et al., 2023; Rosenblum et al., 2020; Slavova et al., 2017; Tran et al., 2021; Zibbell et al., 2019, 2022), while fewer have examined multiple states or the entire US (Jalal & Burke, 2020; Gladden et al., 2016; Marks et al., 2021; McBrien & Alexander, 2022; Sumner et al., 2022; Zibbell et al., 2023; Zoorob et al., 2019). These studies have varied in terms of specific drug seizures examined (e.g., opioid seizures, stimulant seizures), types of drug overdose mortality outcomes (e.g., drug overdose mortality overall, opioid overdose mortality), and units of analysis (e.g., state-year, county-month).

The present study builds on the foundation of the aforementioned studies in several ways. *First*, this study examines associations between state-level drug seizure measures and drug mortality rates not only in the overall US population, but also within subpopulations based on sex, race, and age, in consideration of subpopulation differences in the drugs most frequently involved in overdose deaths (Cano, 2021). *Second*, the study examines the years 2013-2021, as 2013 approximates the beginning of the era of illicitly-manufactured fentanyl (Ciccarone, 2019), and 2021 represents the most recent (finalized, nonprovisional) data available. Prior national studies (e.g., Jalal and Burke, 2020; Sumner et al., 2022; Zibbell et al., 2023) have included data through the year 2019, providing valuable insights yet not capturing more recent trends such as the post-2019 spread of illicitly-manufactured fentanyl beyond the east into the west (Shover et al., 2020) or COVID-era and post-COVID changes in overdose mortality. *Finally*, this study also incorporates measures of xylazine seizures, as xylazine has been identified as an emerging drug threat with a distinct geographic footprint (Kariisa et al., 2023).

Overall, the present study aims to identify the extent to which a variety of measures of drug seizures from the publicly-accessible National Forensic Laboratory Information System account for variation in drug overdose mortality between states and within states over time in various subpopulations of the US. The study uses a national, publicly-available data source to evaluate measures that are accessible to *all* researchers/agencies and available for all states in the US, as many prior studies (e.g., Lowder et al., 2022; Ray et al., 2023; Tran et al., 2021; Zibbell et al., 2019) have utilized state-specific data sources that offer more detailed, in-depth measures (e.g., weekly or monthly data, weights or doses of drugs seized) that are not nationally publicly-available. Findings from the present study may serve as a reference for researchers seeking optimal nationally-available measures of illicit drug availability for statistical controls in evaluations of policy or social/economic context effects on drug overdose, or for public health agencies considering measures to include in early-warning systems (e.g., Hall et al., 2021; Marks et al., 2021; Sumner et al., 2022) for overdose mortality trends.

## 2. Methods

### 2.1 Data Sources and Measures

#### 2.1.1 Independent Variables

Counts of law enforcement drug seizures were obtained from the National Forensic Laboratory Information System (NFLIS-Drug) online Public Data Query System (NFLIS, 2023) as accessed on May 19, 2023. The NFLIS-Drug system compiles data from 286 laboratories, representing approximately 98% of all US drug cases analyzed (Drug Enforcement Administration, n.d.), and each count represents one documented seizure of a specific drug, regardless of the size or weight of the seizure. For each of the 50 states and DC, 2013-2021, we compiled state-by-year counts of seizures positive for: fentanyl-related substances (fentanyl and its analogs); heroin; methamphetamine; cocaine; xylazine; and carfentanil (also included in the fentanyl-related category). We selected these particular substances since they are primarily sold illicitly and are frequently involved in overdose deaths nationwide or in specific regions. We did not examine seizures of prescription opioids, benzodiazepines, or antidepressants, even though these substances are also involved in overdose deaths, since these substances are *both* sourced legally and illicitly, and as such, the extent to which prescription drugs are involved in law enforcement seizures would not likely represent their overall availability (due to not directly accounting for availability via prescription).

We used counts of specific drug seizures to compute *two* different measures for each drug type:

(1) the specific drug seizure *rate* per 1,000 population, comprising the yearly number of law enforcement seizures of the specific drug divided by the population size (using population estimates from the National Center for Health Statistics via the Centers for Disease Control and Prevention [CDC] WONDER online platform; CDC, 2023); and
(2) the specific drug seizure *percentage*, comprising the number of seizures of the specific drug divided by the total summed count of seizures of fentanyl-related substances, heroin, methamphetamine, cocaine, and xylazine.

To provide a specific example, the heroin seizure *rate* in West Virginia, 2020, represents the number of heroin seizures in West Virginia in 2020, divided by the population size of West Virginia in 2020, expressed per 1,000. Alternately, the heroin seizure *percentage* in West Virginia, 2020, represents the number of heroin seizures in West Virginia in 2020, divided by the total (summed) count of seizures of heroin, fentanyl-related substances, methamphetamine, cocaine, and xylazine in West Virginia in 2020, expressed as percent.

#### 2.1.2 Dependent Variables

The primary outcome of interest was the drug overdose mortality rate per 100,000 residents, in each of the 50 states and District of Columbia for each year from 2013 to 2021. Drug overdose mortality rates were obtained from the CDC WONDER online platform (CDC, 2023), representing crude rates for drug overdose deaths of any intent (identified via an underlying cause of death corresponding to International Classification of Disease [ICD]-10 codes X40-44, X60-64, X85, or Y10-14). Overdose deaths involving any drug were the primary outcome of interest, in light of geographic and time variation in the extent to which specific drug types are identified and reported on death certificates (Jones et al., 2019).

Drug overdose mortality rates were examined for the overall resident population as well as for subpopulations identified as: male; female; ages 15-29; ages 30-44; ages 45-74; Non-Hispanic (NH) White; NH Black; and Hispanic. Mortality data suppression requirements resulted in a reduced sample size of 359 state-years for the NH Black population, 335 state-years for the Hispanic population, 451 state-years for the age category 15-29, and 458 state-years for age categories 30-44 and 45-74. Data suppression requirements also limited the study to three broad racial/ethnic groups, as overdose mortality rates were unavailable for other racial/ethnic groups in multiple states. Race-specific drug overdose mortality rates were based on bridged-race categories for years 2013-2020 and single-race categories for the year 2021 (Heron, 2021), in accordance with data availability (CDC, 2023).

### 2.2 Statistical Analyses

Analyses were completed using Stata/MP 18.0, Stata’s user-written program heatplot (Jann, 2019), and RStudio. We calculated descriptive statistics (minimum and maximum values, means, and standard deviations) for all measures used in the study. Next, we examined between-state variation in drug overdose mortality and drug seizures, calculating Pearson correlation coefficients for states’ levels of different types of drug seizures and drug overdose mortality rates, year by year. We calculated the correlations for each year individually due to the non-independence of states’ observations over time and the potential for correlations between different drug seizure measures and overdose mortality rates to shift over the years examined.

For the study’s main analyses, we used two-way (state and year) fixed-effects regression models (using Stata’s *xtreg, fe* with year dummy variables) to predict log-transformed drug overdose mortality rates, overall and by subpopulation (sex, age, and racial/ethnic group), using panel data from 2013-2021 and robust standard errors clustered at the state level. Models comprised:

A. Baseline models with no predictors beyond the state and year fixed effects;
B. Models adding one drug seizure measure per model; and
C. Models adding all drug seizure *rates* simultaneously *or* all drug seizure *percentages* simultaneously (with the carfentanil measure omitted, since already included in the fentanyl-related category, and one additional category [heroin] omitted to avoid perfect collinearity).

In these models, all predictors (drug seizure measures) were standardized with a mean of zero and standard deviation of one. For each model, we report within-R-squared values (Allison, 2009) and Akaike Information Criterion (AIC) values for assessment of model fit, with lower AIC values within each set of models indicating greater predictive accuracy, while accounting for the number of parameters estimated (Cavanaugh et al., 2019). AIC values were evaluated due to their simulation-based validation with fixed effects regression models for panel data (Yum, 2022).

## 3. Results

Summary statistics for all measures in the study are presented in Table 1, and Figure 1 depicts the percentage of US drug seizures each year corresponding to the drug types examined in the study. As presented in Figure 1, in the US overall, the proportional representation of cocaine and heroin (in the drug seizures examined) decreased over time (2013-2021), while seizures of fentanyl-related substances, methamphetamine, and xylazine increased in relative representation over time. In Supplemental Material, Figures S1-S6 display state-year seizure rates and percentages for each drug examined in the study, highlighting differences between states in drug seizure trends. In many states, drug seizure measures in the form of rates per population followed similar trends as measures in the form of percentages of total seizures, while in other states, time trends in these two types of measures appeared to differ notably.

**Figure 1.**
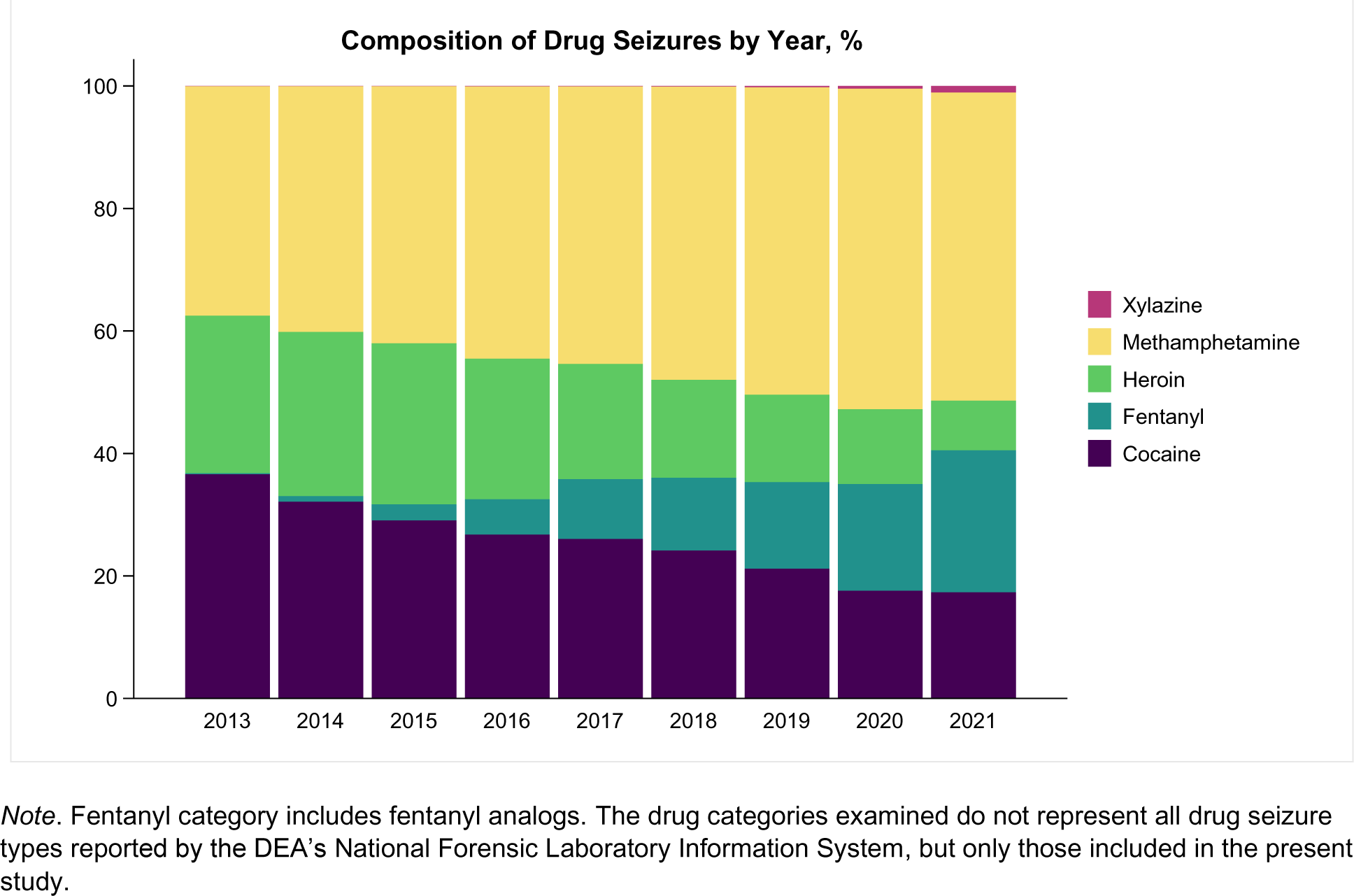
Percentage of drug seizures each year (United States, 2013-2021) corresponding to each of the drug types examined in the study.

**Table 1.** Descriptive Characteristics of the State-Level Measures Included in the Study, United States, 2013-2021.

|  | Mean (SD) | Min, Max | SD, between | SD, within |
| --- | --- | --- | --- | --- |
| **Drug overdose mortality rates, per 100,000 population:** |  |  |  |  |
| for US, overall | 21.95 (11.12) | 2.76, 84.19 | 8.60 | 7.15 |
| for male population | 29.18 (16.64) | 3.24, 115.40 | 12.76 | 10.82 |
| for female population | 14.99 (6.28) | 4.81, 53.14 | 5.04 | 3.81 |
| for NH White population | 23.94 (11.38) | 3.13, 85.54 | 9.35 | 6.61 |
| for NH Black population | 25.52 (21.00) | 0.00, 137.36 | 15.60 | 15.36 |
| for Hispanic population | 13.03 (9.18) | 0.00, 54.66 | 7.89 | 5.76 |
| for population aged 15-29 | 19.49 (9.92) | 3.35, 59.88 | 7.59 | 6.45 |
| for population aged 30-44 | 41.92 (25.75) | 7.28, 206.21 | 20.55 | 15.76 |
| for population aged 45-74 | 28.00 (16.70) | 5.08, 176.42 | 13.60 | 9.88 |
| **Law enforcement drug seizure measures:** |  |  |  |  |
| Fentanyl seizure rate | 0.26 (0.45) | 0.00, 2.89 | 0.32 | 0.33 |
| Fentanyl % | 9.53 (13.13) | 0.00, 60.56 | 8.84 | 9.78 |
| Heroin seizure rate | 0.44 (0.45) | 0.00, 3.15 | 0.38 | 0.24 |
| Heroin % | 19.86 (15.99) | 0.30, 81.60 | 12.34 | 10.31 |
| Methamphetamine seizure rate | 1.26 (1.20) | 0.00, 6.07 | 1.10 | 0.49 |
| Methamphetamine % | 47.36 (29.38) | 0.34, 96.41 | 28.75 | 7.17 |
| Cocaine seizure rate | 0.49 (0.40) | 0.01, 2.16 | 0.37 | 0.15 |
| Cocaine % | 22.95 (16.30) | 2.06, 80.92 | 14.65 | 7.41 |
| Xylazine seizure rate | 0.01 (0.02) | 0.00, 0.28 | 0.01 | 0.02 |
| Xylazine % | 0.30 (1.32) | 0.00, 17.48 | 0.66 | 1.15 |
| Carfentanil seizure rate | 0.00 (0.02) | 0.00, 0.39 | 0.01 | 0.02 |
| Carfentanil % | 0.07 (0.36) | 0.00, 5.98 | 0.20 | 0.30 |

Notes. Based on publicly available data from the US National Forensic Laboratory Information System, Public Data Query System (Retrieved May 19, 2023) for the 50 states and District of Columbia. Drug seizure rates are expressed per 1,000 population. *Abbreviations.* %, percentage; SD, standard deviation; SD between, standard deviation between states; SD within, standard deviation within states over time.

### 3.1 Variation Between States

Figure 2 presents yearly Pearson correlation coefficients for state levels of various drug seizure measures and drug overdose mortality rates, with darker colors indicating stronger correlations (positive correlations in red and negative correlations in blue). For nearly every drug type examined, stronger correlations were observed between drug overdose mortality rates and measures corresponding to drug seizure *percentages*, rather than drug seizure *rates per population*. Correlations differed over time; for example, the correlation between state drug overdose mortality rates and heroin seizure percentage declined over time, from a peak of Pearson’s *r* = 0.56 in 2015 to a correlation near zero in the years 2020 and 2021. For most other drug seizure measures besides heroin, correlations with state drug overdose mortality rates peaked during the years 2016-2019 and were lower in the two most recent years examined (2020 and 2021). Overall, the measure exhibiting the strongest correlation with state-level drug overdose mortality rates in any given year was the methamphetamine seizure percentage in the year 2018, negatively correlated (*r* = -0.72) with drug overdose mortality rates in the year 2018, followed by the fentanyl-related seizure percentage in the years 2018 and 2019, positively correlated (*r* = 0.71) with 2018 and 2019 drug overdose mortality rates, respectively.

**Figure 2.**
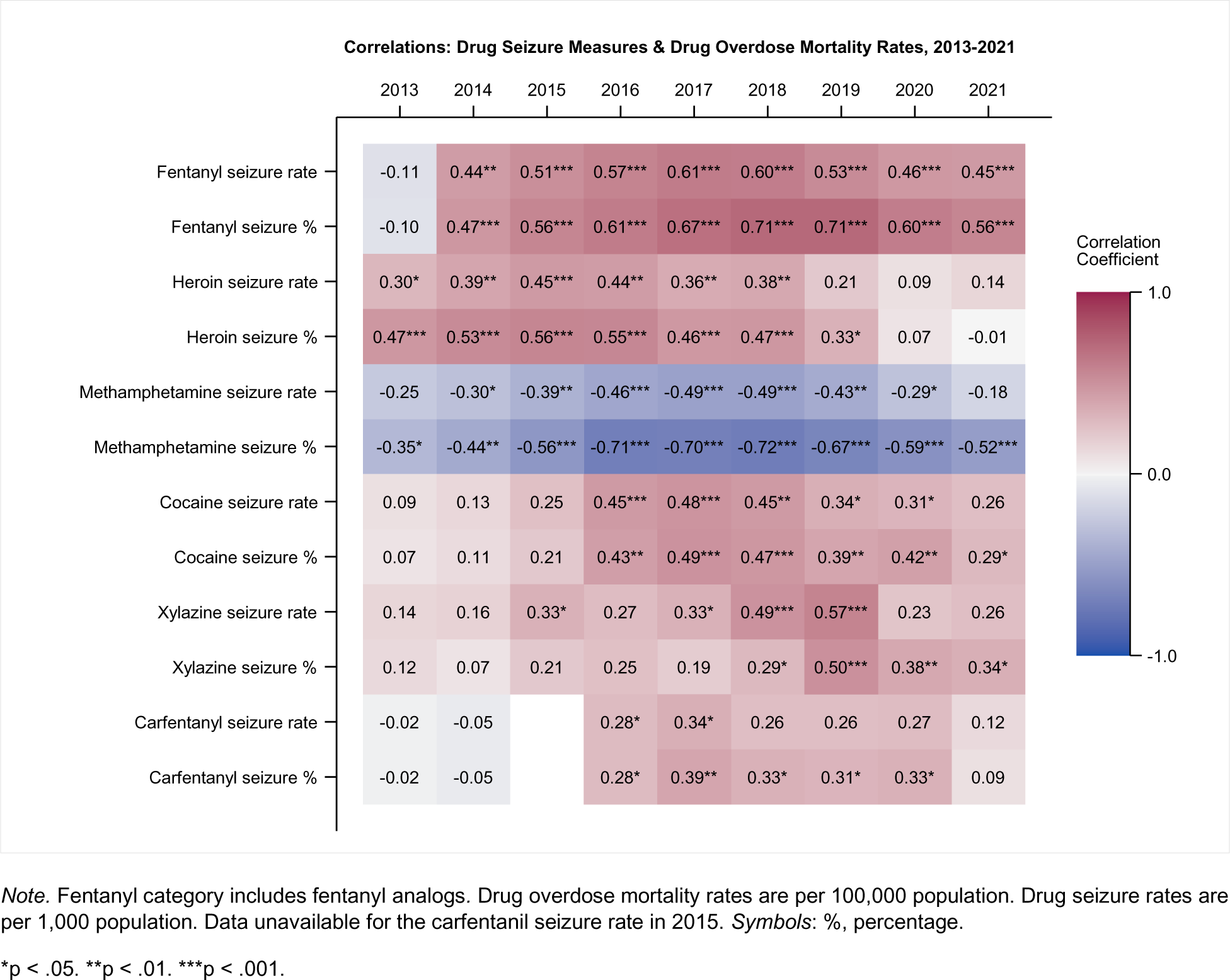
Pearson Correlation Coefficients for Drug Seizure Measures and Annual Drug Overdose Mortality Rates, United States, 2013-2021.

### 3.2 Variation Within States Over Time

Tables 2-4 provide results from state- and year-fixed effects regression models predicting state-level drug overdose mortality rates by various drug seizure measures. Table 2 includes results in the overall population and among male and female populations, while Table 3 presents results by racial/ethnic subgroup, and Table 4 provides results by age group.

**Table 2.** Results of state- and year-fixed effects regression models predicting drug overdose mortality rates in the US overall, and in male and female populations, by drug seizure measures, 2013-2021 (*n*=459 state-years)

|  | **Overall** |  |  | **Male Population** |  |  | **Female Population** |  |  |
| --- | --- | --- | --- | --- | --- | --- | --- | --- | --- |
| | $\beta$ (SE) | R<sup>2</sup> | AIC | $\beta$ (SE) | R<sup>2</sup> | AIC | $\beta$ (SE) | R<sup>2</sup> | AIC |
| **A. Baseline Models** |  |  |  |  |  |  |  |  |  |
| State & Year Fixed Effects |  | 0.73 | -448.75 |  | 0.78 | -413.26 |  | 0.56 | -443.69 |
| **B. Baseline Models + One Drug Seizure Measure at a Time** |  |  |  |  |  |  |  |  |  |
| Fentanyl-related seizure rate | **0.08** (0.02) | 0.76 | -491.39 | **0.07** (0.02) | 0.79 | -439.11 | **0.09** (0.02) | 0.62 | -505.99 |
| Fentanyl-related seizure percentage | **0.14** (0.02) | 0.79 | -552.96 | **0.12** (0.02) | 0.81 | -482.39 | **0.15** (0.02) | 0.68 | -578.91 |
| Heroin seizure rate | -0.03 (0.02) | 0.73 | -450.62 | -0.00 (0.02) | 0.78 | -411.27 | -0.07 (0.02) | 0.59 | -466.52 |
| Heroin seizure percentage | **-0.07** (0.02) | 0.75 | -477.40 | **-0.04** (0.02) | 0.78 | -421.13 | **-0.11** (0.02) | 0.63 | -520.57 |
| Meth. seizure rate | -0.04 (0.04) | 0.73 | -450.02 | -0.04 (0.04) | 0.78 | -414.55 | -0.03 (0.04) | 0.57 | -444.14 |
| Meth. seizure percentage | 0.05 (0.06) | 0.73 | -448.94 | 0.01 (0.06) | 0.78 | -411.29 | **0.14** (0.06) | 0.58 | -459.47 |
| Cocaine seizure rate | -0.00 (0.03) | 0.73 | -446.76 | -0.01 (0.03) | 0.78 | -411.43 | 0.01 (0.03) | 0.56 | -441.78 |
| Cocaine seizure percentage | **-0.11** (0.04) | 0.75 | -475.46 | **-0.12** (0.04) | 0.79 | -439.55 | **-0.12** (0.04) | 0.59 | -471.09 |
| Xylazine seizure rate | -0.01 (0.01) | 0.73 | -447.38 | **-0.02** (0.01) | 0.78 | -415.10 | 0.01 (0.01) | 0.57 | -443.03 |
| Xylazine seizure percentage | 0.01 (0.01) | 0.73 | -448.29 | 0.00 (0.01) | 0.78 | -411.27 | **0.03** (0.01) | 0.57 | -450.38 |
| Carfentanil seizure rate | **0.02** (0.00) | 0.73 | -451.62 | **0.02** (0.00) | 0.78 | -415.31 | **0.02** (0.00) | 0.57 | -447.08 |
| Carfentanil seizure percentage | **0.02** (0.01) | 0.73 | -453.47 | **0.02** (0.01) | 0.78 | -416.94 | **0.02** (0.00) | 0.57 | -449.19 |
| **C. Baseline Models + All Drug Seizure Measures Simultaneously** |  |  |  |  |  |  |  |  |  |
| All drug seizure rates<sup>a</sup> |  | 0.77 | -519.08 |  | 0.81 | -482.66 |  | 0.63 | -513.82 |
| All drug seizure percentages<sup>ab</sup> |  | 0.80 | -580.23 |  | 0.83 | -520.23 |  | 0.70 | -606.95 |

Notes. <sup>a</sup>Excluding carfentanil seizures, which are already included in the fentanyl-related category. <sup>b</sup>Also excluding one category (heroin) to avoid perfect collinearity. Seizure *rate* refers to the number of law enforcement seizures of the specified drug per 1,000 population. Seizure *percentage* refers to the number of seizures of the specified drug divided by all drug seizures examined, expressed as percent. Drug overdose mortality rates are per 100,000 population and are log-transformed. All predictors are standardized with a mean of 0 and standard deviation of 1. Robust standard errors are clustered at the state level. Coefficients in bold are significantly different from zero at $p < 0.05$ . R<sup>2</sup> is for the within variation. *Abbreviations.* SE, standard error; AIC, Akaike information criterion; Meth, methamphetamine.

**Table 3.** Results of state- and year-fixed effects regression models predicting drug overdose mortality rates for Non-Hispanic (NH) White, NH Black, and Hispanic populations, by drug seizure measures, 2013-2021.

|  | **White, Non-Hispanic** |  |  | **Black, Non-Hispanic** |  |  | **Hispanic** |  |  |
| --- | --- | --- | --- | --- | --- | --- | --- | --- | --- |
| | $\beta$ (SE) | R<sup>2</sup> | AIC | $\beta$ (SE) | R<sup>2</sup> | AIC | $\beta$ (SE) | R<sup>2</sup> | AIC |
| **A. Baseline Models** |  |  |  |  |  |  |  |  |  |
| State & Year Fixed Effects |  | 0.65 | -510.57 |  | 0.89 | -232.75 |  | 0.74 | -58.19 |
| **B. Baseline Models + One Drug Seizure Measure at a Time** |  |  |  |  |  |  |  |  |  |
| Fentanyl-related seizure rate | **0.09** (0.02) | 0.71 | -575.95 | **0.06** (0.03) | 0.90 | -242.42 | **0.11** (0.03) | 0.76 | -79.33 |
| Fentanyl-related seizure percentage | **0.13** (0.02) | 0.73 | -621.60 | **0.10** (0.03) | 0.90 | -256.74 | **0.16** (0.04) | 0.77 | -96.04 |
| Heroin seizure rate | -0.03 (0.02) | 0.66 | -513.87 | -0.02 (0.03) | 0.89 | -231.96 | -0.03 (0.05) | 0.74 | -57.50 |
| Heroin percentage | **-0.08** (0.02) | 0.69 | -551.42 | -0.05 (0.03) | 0.89 | -237.56 | -0.06 (0.05) | 0.74 | -61.10 |
| Meth seizure rate | -0.01 (0.03) | 0.66 | -508.77 | 0.01 (0.05) | 0.89 | -230.98 | 0.04 (0.07) | 0.74 | -56.96 |
| Meth percentage | 0.06 (0.06) | 0.66 | -512.81 | 0.06 (0.06) | 0.89 | -232.85 | 0.04 (0.10) | 0.74 | -56.69 |
| Cocaine seizure rate | 0.03 (0.03) | 0.66 | -511.88 | -0.00 (0.04) | 0.89 | -230.76 | 0.03 (0.06) | 0.74 | -56.72 |
| Cocaine percentage | **-0.09** (0.04) | 0.68 | -530.02 | **-0.10** (0.04) | 0.90 | -243.15 | **-0.22** (0.07) | 0.76 | -85.78 |
| Xylazine seizure rate | -0.01 (0.01) | 0.66 | -509.88 | -0.02 (0.01) | 0.89 | -233.82 | -0.01 (0.01) | 0.74 | -56.37 |
| Xylazine percentage | 0.00 (0.01) | 0.66 | -508.67 | -0.00 (0.01) | 0.89 | -230.81 | 0.00 (0.01) | 0.74 | -56.20 |
| Carfentanil seizure rate | **0.02** (0.00) | 0.66 | -514.65 | **0.02** (0.00) | 0.89 | -233.62 | **0.02** (0.00) | 0.74 | -58.34 |
| Carfentanil percentage | **0.02** (0.00) | 0.67 | -516.93 | **0.02** (0.01) | 0.89 | -234.92 | **0.02** (0.01) | 0.74 | -58.93 |
| **C. Baseline Models + All Drug Seizure Measures Simultaneously** |  |  |  |  |  |  |  |  |  |
| All drug seizure rates<sup>a</sup> |  | 0.72 | -592.25 |  | 0.90 | -244.73 |  | 0.76 | -78.76 |
| All drug seizure percentages<sup>ab</sup> |  | 0.75 | -641.18 |  | 0.90 | -267.26 |  | 0.79 | -121.36 |

Notes. <sup>a</sup>Excluding carfentanil seizures, which are already included in the fentanyl-related category. <sup>b</sup>Also excluding one category (heroin) to avoid perfect collinearity. State-year observations: NH White, $n=459$ ; NH Black, $n=359$ ; Hispanic, $n=335$ . Seizure *rate* refers to the number of law enforcement seizures of the specified drug per 1,000 population. Seizure *percentage* refers to the number of seizures of the specified drug divided by all drug seizures examined, expressed as percent. Drug overdose mortality rates are per 100,000 population and are log-transformed. All predictors are standardized with a mean of 0 and standard deviation of 1. Robust standard errors are clustered at the state level. Coefficients in bold are significantly different from zero at $p<0.05$ . R<sup>2</sup> is for the within variation. Abbreviations. NH, Non-Hispanic; SE, standard error; AIC, Akaike information criterion; Meth, methamphetamine.

**Table 4.** Results of state- and year-fixed effects regression models predicting drug overdose mortality rates for populations age 15-29, 30-44, and 45-74, by drug seizure measures, 2013-2021.

|  | <u>Ages 15-29</u> |  |  | <u>Ages 30-44</u> |  |  | <u>Ages 45-74</u> |  |  |
| --- | --- | --- | --- | --- | --- | --- | --- | --- | --- |
| | $\beta$ (SE) | R<sup>2</sup> | AIC | $\beta$ (SE) | R<sup>2</sup> | AIC | $\beta$ (SE) | R<sup>2</sup> | AIC |
| **A. Baseline Models** |  |  |  |  |  |  |  |  |  |
| State & Year<br>Fixed Effects |  | 0.60 | -172.70 |  | 0.71 | -283.72 |  | 0.64 | -429.08 |
| **B. Baseline Models + One Drug Seizure Measure at a Time** |  |  |  |  |  |  |  |  |  |
| Fentanyl-related<br>seizure rate | **0.06** (0.02) | 0.61 | -184.29 | **0.09** (0.02) | 0.73 | -325.31 | **0.08** (0.02) | 0.68 | -472.93 |
| Fentanyl-related<br>seizure<br>percentage | **0.08** (0.03) | 0.61 | -188.17 | **0.17** (0.02) | 0.77 | -396.82 | **0.15** (0.03) | 0.73 | -543.53 |
| Heroin seizure<br>rate | **0.06** (0.03) | 0.61 | -181.57 | -0.04 (0.02) | 0.71 | -287.33 | **-0.07** (0.02) | 0.67 | -453.45 |
| Heroin<br>percentage | 0.02 (0.03) | 0.60 | -172.53 | **-0.09** (0.02) | 0.73 | -318.23 | **-0.11** (0.02) | 0.70 | -500.65 |
| Meth seizure<br>rate | -0.03 (0.04) | 0.60 | -171.64 | -0.04 (0.04) | 0.71 | -284.73 | -0.01 (0.05) | 0.65 | -427.48 |
| Meth<br>percentage | -0.10 (0.08) | 0.60 | -176.13 | **0.13** (0.06) | 0.71 | -292.71 | **0.13** (0.05) | 0.66 | -442.79 |
| Cocaine<br>seizure rate | 0.05 (0.03) | 0.60 | -173.96 | -0.01 (0.03) | 0.71 | -281.95 | -0.03 (0.03) | 0.65 | -428.57 |
| Cocaine<br>percentage | **-0.11** (0.05) | 0.61 | -185.56 | **-0.18** (0.04) | 0.74 | -334.03 | **-0.11** (0.04) | 0.66 | -450.57 |
| Xylazine<br>seizure rate | **-0.04** (0.01) | 0.61 | -187.04 | -0.01 (0.01) | 0.71 | -282.90 | 0.02 (0.01) | 0.65 | -430.47 |
| Xylazine<br>percentage | **-0.04** (0.01) | 0.61 | -185.21 | 0.01 (0.01) | 0.71 | -282.27 | **0.04** (0.01) | 0.66 | -444.15 |
| Carfentanil<br>seizure rate | **0.02** (0.00) | 0.60 | -173.14 | **0.02** (0.00) | 0.71 | -286.68 | **0.02** (0.00) | 0.65 | -430.56 |
| Carfentanil<br>percentage | **0.02** (0.01) | 0.60 | -174.96 | **0.03** (0.01) | 0.71 | -289.02 | **0.02** (0.00) | 0.65 | -431.30 |
| **C. Baseline Models + All Drug Seizure Measures Simultaneously** |  |  |  |  |  |  |  |  |  |
| All drug seizure<br>rates<sup>a</sup> |  | 0.67 | -250.43 |  | 0.75 | -353.31 |  | 0.69 | -481.35 |
| All drug seizure<br>percentages<sup>ab</sup> |  | 0.66 | -247.03 |  | 0.80 | -450.90 |  | 0.74 | -567.37 |

Notes. <sup>a</sup>Excluding carfentanil seizures, which are already included in the fentanyl-related category. <sup>b</sup>Also excluding one category (heroin) to avoid perfect collinearity. State-year observations: Ages 15-29, $n=451$ ; Ages 30-44, $n=458$ ; Ages 45-74, $n=458$ . Seizure *rate* refers to the number of law enforcement seizures of the specified drug per 1,000 population. Seizure *percentage* refers to the number of seizures of the specified drug divided by all drug seizures examined, expressed as percent. Drug overdose mortality rates are per 100,000 population and are log-transformed. All predictors are standardized with a mean of 0 and standard deviation of 1. Robust standard errors are clustered at the state level. Coefficients in bold are significantly different from zero at $p<0.05$ . R<sup>2</sup> is for the within variation. *Abbreviations.* SE, standard error; AIC, Akaike information criterion; Meth, methamphetamine.

In models examining one drug seizure measure per model, the fentanyl-related seizure percentage (and, to a lower extent, the fentanyl-related seizure rate and the carfentanil seizure rate and percentage) was significantly positively associated with drug overdose mortality rates across all subgroups examined, while the cocaine seizure percentage was significantly negatively associated with drug overdose mortality rates across all subgroups. The direction and significance of associations with other drug seizure measures differed between subgroups; for example, the heroin seizure percentage was significantly negatively associated with drug overdose mortality rates for all subgroups except ages 15-29, NH Black, and Hispanic.

The within R-squared and AIC values for baseline models (with state and year fixed effects only) varied considerably between subgroups examined, and, similarly, the extent to which the addition of drug seizure measures increased within R-squared values also differed between subpopulations. For example, in the female subpopulation, the model with all drug seizure percentage measures accounted for 70% of the within-variation in the outcome, compared to 56% in the baseline model; in contrast, the model with all drug seizure percentage measures accounted for 90% of the within-variation in the outcome for the NH Black subpopulation, relatively similar to the 89% in the baseline model.

For the overall population and all subgroups examined, except for ages 15-29, the model that included *all* drug seizure *percentage* measures represented the best-performing model examined (as identified via the lowest AIC and highest within R-squared values), followed by the model including only the fentanyl-related seizure *percentage*, although in the male subpopulation the AIC was relatively similar in the model with all drug seizure rates (−482.66) and the model with the fentanyl-related seizure percentage only (−482.39). In contrast, in the population age 15-29, the best-performing model included all drug seizure *rates* (AIC -250.43), closely followed by the model with all drug seizure *percentages* (AIC -247.03).

## 4. Limitations

Although the drug seizure data source used in the present study (NFLIS) is publicly-accessible, comparable between years, and available for all states (Drug Enforcement Administration, n.d.), it does not include the level of detail available in state-specific data sources examined in prior studies, such as weights/quantities/doses within each drug seizure, the week or month during which the drug seizure occurred, the county, zip code area, or address corresponding to the seizure, combinations of drugs seized, or the type of law enforcement agency involved. While the NFLIS contains data from an estimated 98% of all US drug cases, not all forensic laboratories are represented in the NFLIS, not drugs seized are analyzed, laboratories may differ in terms of which drugs are analyzed and reported, and exact counts of seizures depend on the day results were retrieved, due to continuous updates of delayed data (Drug Enforcement Administration, n.d.).

The state- and year-fixed effects models used in the present study included a variety of drug seizure measures, yet not all drug seizure types were included, and models did not account for availability of prescription drugs, given the present study’s focus on illicit drugs. Moreover, drug seizure counts do not differentiate between illicitly-manufactured fentanyl and diverted pharmaceutical fentanyl or between seizures of different quantities or weights. Finally, in preliminary analyses examining variation between states, correlation coefficients were computed for each year independently in consideration of temporal, yet not spatial, dependence of observations.

## 5. Discussion

The present study examined the extent to which publicly-accessible law enforcement drug seizure measures account for differences in overall drug overdose mortality rates between states, within states over time, and in various subpopulations. Drug seizure measures were conceptualized as proxies for street drug availability, consistent with numerous prior studies (e.g., Jalal and Burke, 2021; McBrien and Alexander, 2022; Zibbell et al., 2019, 2023; Zoorob, 2019), including several by the CDC (e.g., Gladden et al., 2016; Sumner et al., 2022). In contrast, several studies (e.g., Lowder et al., 2022; Mohler et al., 2021; Ray et al., 2023) have examined drug seizures as law enforcement operations with the potential to either reduce drug overdose deaths (through limiting the availability and use of drugs) or increase drug overdoses (through shifting individuals who use drugs to less familiar drug sources and products of unknown potency). The present study, however, was not based on the hypothesis that drug seizures directly impact rates of drug overdose mortality, but rather that higher proportional representation of potent substances (e.g., fentanyl, carfentanil) in drug seizure data is indicative of greater availability of these substances, accompanied by more use of these substances (relative to others), and a higher rate of fatal overdoses.

In a recent analysis of 2014-2019 US opioid overdose deaths by state and quarter, drug seizure measures expressed as *counts* were more strongly associated with changes in opioid overdose deaths, compared to drug seizure measures expressed as *percentages* (Zibbell et al., 2023). In contrast, in the present study’s examination of 2013-2021 rates of drug overdose mortality, drug seizure measures expressed as *percentages* of total seizures accounted for more variation in drug overdose mortality between and within states (in most, yet not all models), relative to measures expressed as drug seizure *rates* per population. It is plausible that drug seizure rates per population may be more sensitive to year-by-year or between-state differences in overall levels of drug enforcement, while percentage-based measures may more stably reflect the relative availability of one substance compared to another, regardless of changes in total numbers of drug seizures (Zibbell et al., 2023). With the exception of a few studies that have included percentage-based measures (Zibbell et al., 2019, 2023) the majority of studies on associations between drug seizures and overdose mortality have examined counts or rates of drug seizures (Gladden et al., 2016; Hall et al., 2021; Jalal and Burke, 2020; Lowder et al., 2022; Rosenblum et al., 2020; Slavova et al., 2017; Sumner et al., 2022; Tran et al., 2021; Zibbell et al., 2022; Zoorob, 2019). Findings from the present study, however, support the utility of proportion-based drug seizure measures in predicting state-level rates of drug overdose mortality.

Of the state- and year-fixed effects models examined in this study, the best-performing model for the overall population (and nearly all subgroups examined) included percentage-based measures for seizures involving fentanyl-related substances, cocaine, methamphetamine, and xylazine (with the percentage involving heroin omitted to avoid perfect collinearity). When examining only one drug-seizure measure at a time, models with the *fentanyl*-related seizure percentage accounted for the largest extent of variation in state-level drug overdose mortality rates. Fentanyl’s high potency, wide availability (Kilmer et al., 2022), and involvement in an estimated two thirds of all drug overdose deaths (Spencer et al., 2023) may explain the strong performance of fentanyl-related seizure measures as predictors of overall drug overdose mortality rates. The results of multiple prior studies support the positive association between fentanyl-related seizures and drug overdose deaths in individual states and the US overall (Gladden et al., 2016; Rosenblum et al., 2020; Slavova et al., 2017; Zibbell et al., 2019, 2023; Zoorob, 2019), and results of the present study add that this association is observed across subpopulations based on sex, race/ethnicity, and age.

With respect to specific fentanyl analogs of even higher potency than fentanyl, the present study also documented significant positive associations between state-level carfentanil seizures and drug overdose mortality rates, yet carfentanil seizures accounted for a lower proportion of the variance in drug overdose mortality rates, compared to other drug seizure measures. These results are consistent with a 2014-2018 Ohio analysis indicating a stronger association between drug overdose deaths and fentanyl-related seizures than carfentanil seizures specifically (Tran et al., 2021), while contrasting with a nationwide analysis attributing the 2017-2018 growth in drug overdose mortality to increases in carfentanil seizure rates (Jalal and Burke, 2020). Despite carfentanil’s high level of potency, its availability within only a few states, and only during a few years (as depicted in Supplemental Figure S6), likely diminishes the association between carfentanil and overall overdose mortality across the US over multiple years of data.

State-level correlations between drug seizure measures and drug overdose mortality rates differed over the years examined in the study (2013-2021). The attenuation over time of the state-level correlation between heroin seizure measures and drug overdose mortality rates is consistent with documented trends in drug availability and overdose deaths, as heroin has diminished in availability and in involvement in overdose deaths over time, virtually replaced by fentanyl (Pardo et al., 2021). The relatively weaker correlations between most drug seizure measures and drug overdose mortality rates in the years 2020-2021, compared to 2019, may also possibly reflect COVID-19-related disruptions in drug markets or increased contributions of social factors to drug overdose mortality during the unique circumstances of the pandemic (Imtiaz et al., 2021). Nonetheless, further research would be necessary to test these potential explanations or to clarify post-COVID trends as more recent data become available.

## 6. Conclusions and Implications

While researchers have highlighted the role of drug supplies (Ruhm, 2019) in shaping risk of fatal overdose, many have also highlighted how a sole focus on supply-side interventions, such as policies aimed at reducing prescription opioid access without simultaneously addressing addiction and underlying causes of substance use, can lead to increases in the availability and use of more dangerous and potent drugs, ultimately increasing deaths (Beletsky and Davis, 2017; Lowder et al., 2022; McClean et al., 2022; Rosenblum et al., 2020). Although examining drug seizure measures may emphasize *supply*-side influences on drug overdose mortality, drug seizure measures can also facilitate evaluations of *demand*-side policies (e.g., naloxone access laws, Good Samaritan laws, cannabis legalization; McClean et al., 2022) and social/economic influences (e.g., unemployment, poverty, incarceration), since “isolating the effects” of such policies and socioeconomic factors may require statistical adjustment for time-varying supply-side factors such as drug availability. Overall, findings of the present study support the potential utility of publicly-available drug seizure proportional measures (i.e., percentages of drug seizures involving various drug types), especially the proportion of fentanyl-related seizures, for overdose early warning systems (e.g., Hall et al., 2021; Marks et al., 2021; Sumner et al., 2022) and research into contextual-level risk factors and interventions, with the overarching aim of reducing the number of lives lost to drug overdoses each year in the US.

## Data Availability

All data are available online at
- National Center for Health Statistics, Mortality Data on CDC WONDER (https://wonder.cdc.gov/mcd.html)
- National Forensic Laboratory Information System, Drug Enforcement Administration (https://www.nflis.deadiversion.usdoj.gov/)

https://wonder.cdc.gov/mcd.html

https://www.nflis.deadiversion.usdoj.gov/

## Supplemental Material

**Figure S1.**
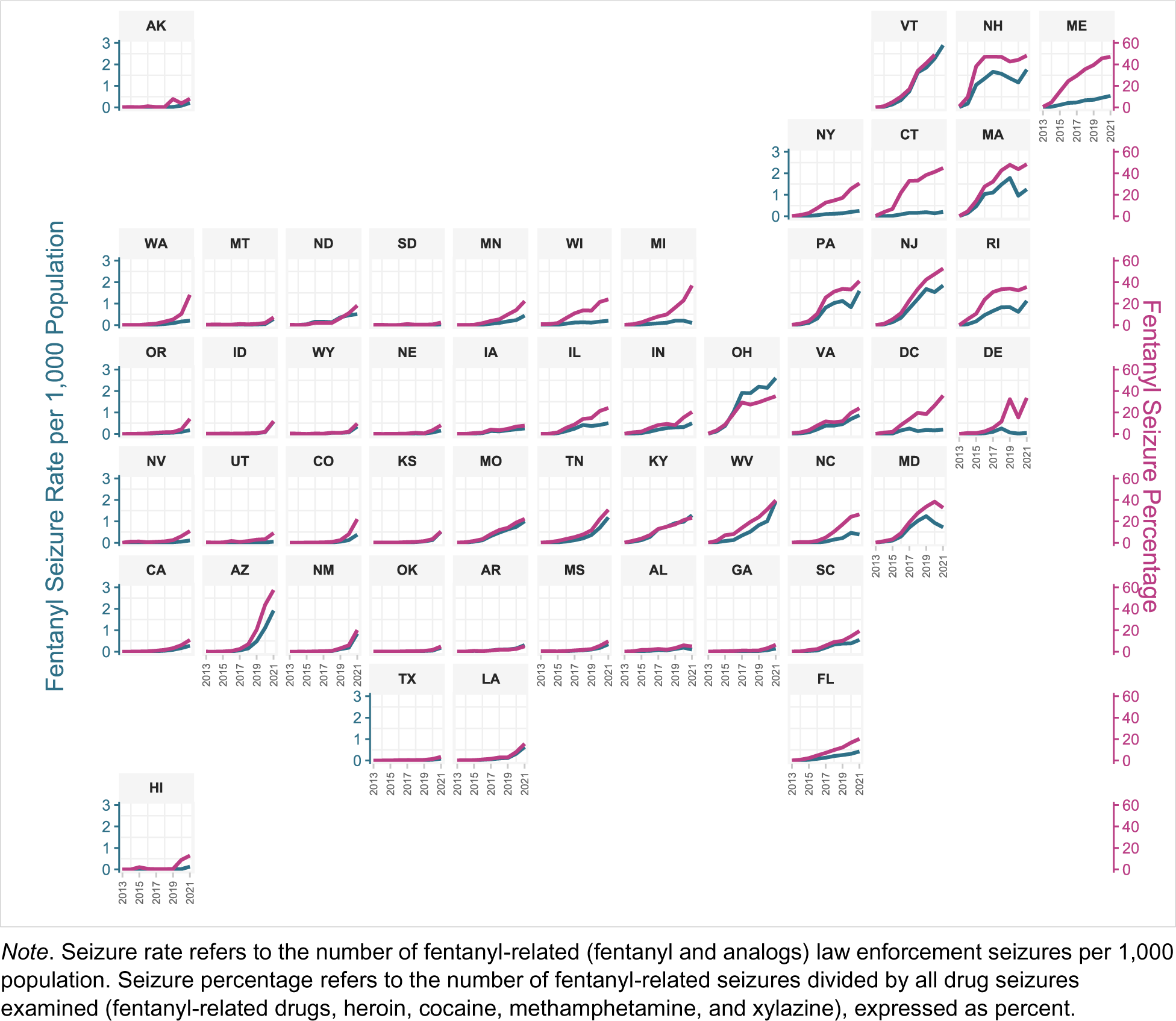
Distributions of Fentanyl-Related Seizure Rates/Percentages by State and Year, US, 2013-2021.

**Figure S2.**
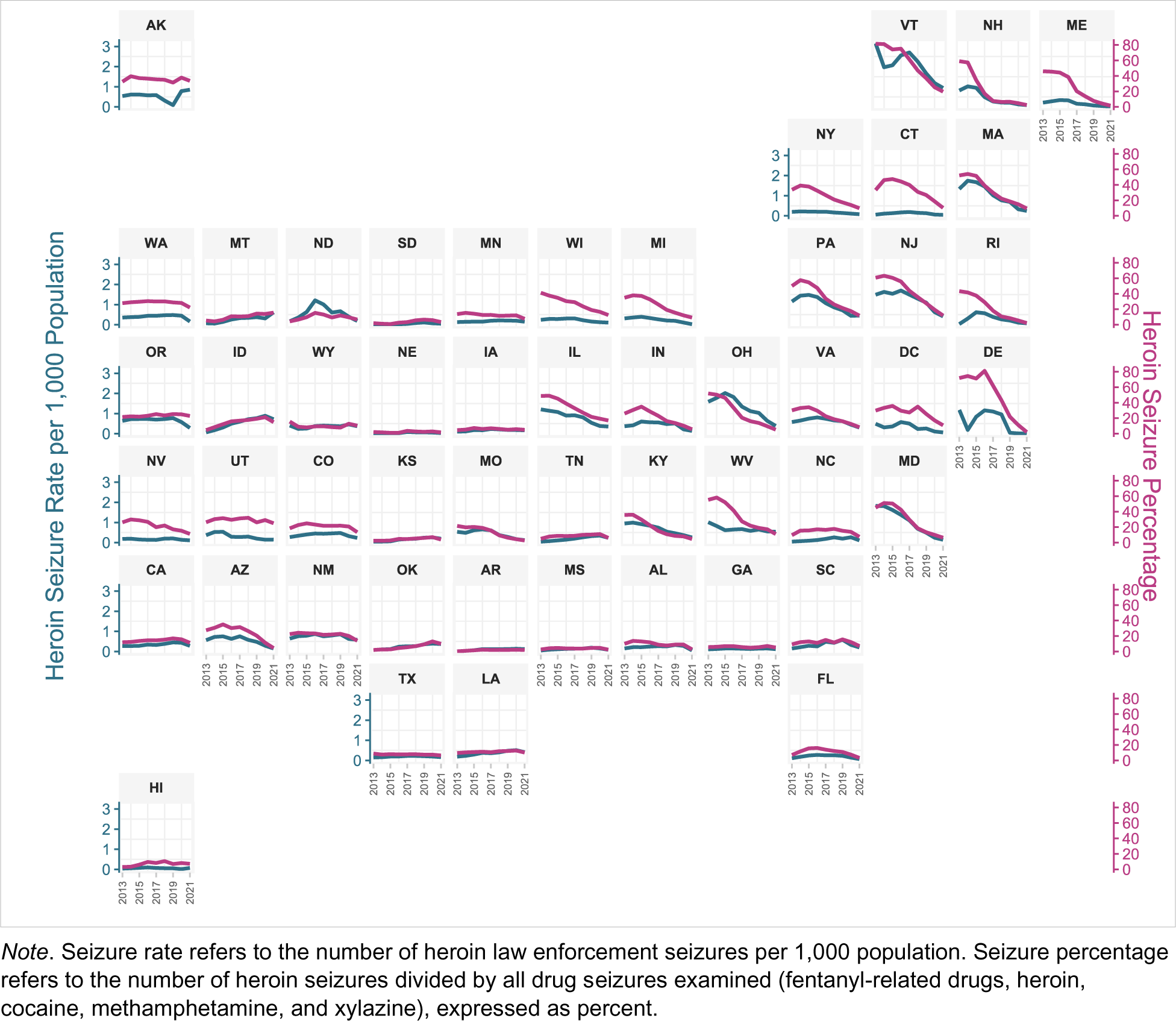
Distributions of Heroin Seizure Rates/Percentages by State, US, 2013-2021.

**Figure S3.**
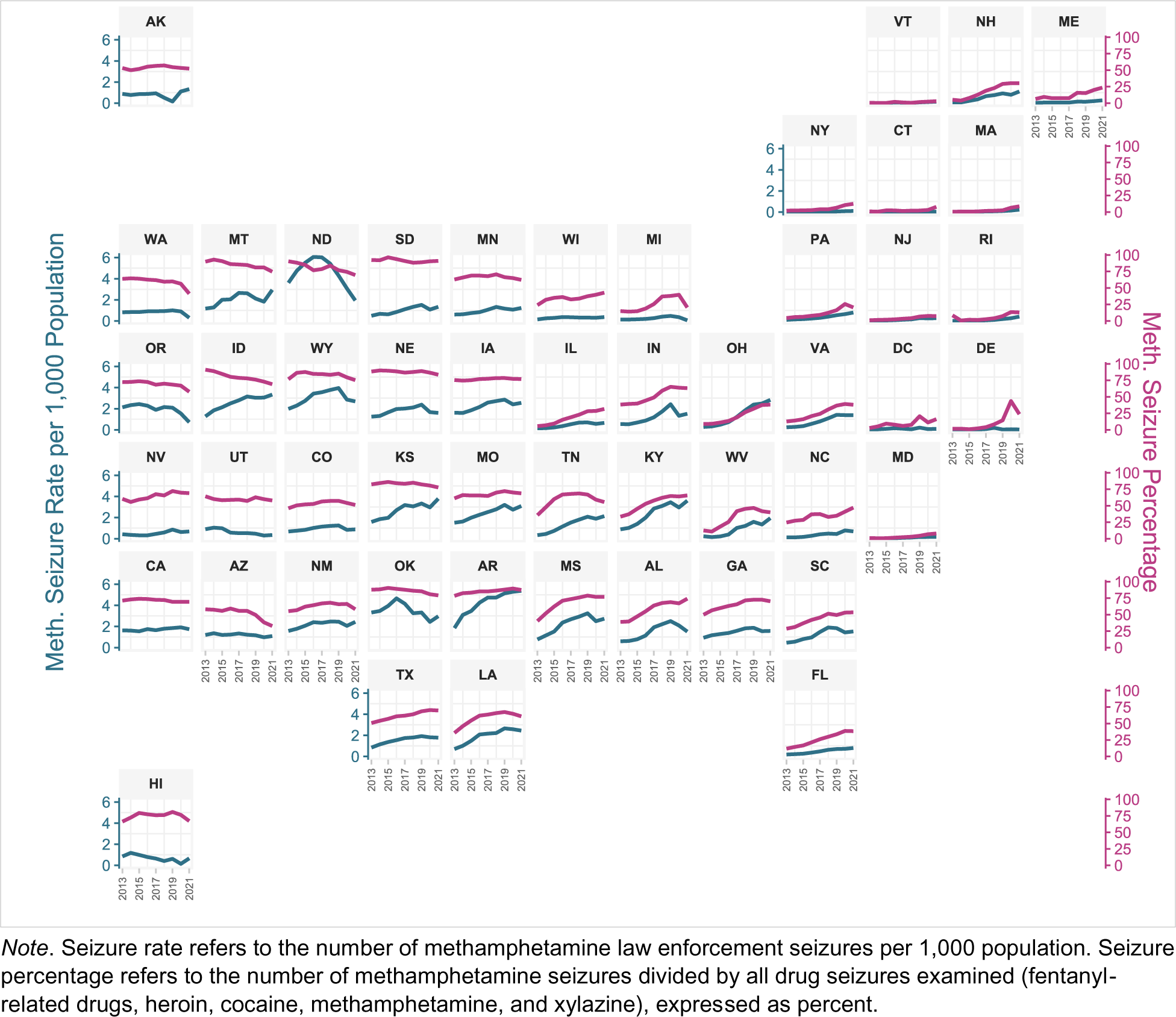
Distributions of Methamphetamine Seizure Rates/Percentages by State, US, 2013-2021.

**Figure S4.**
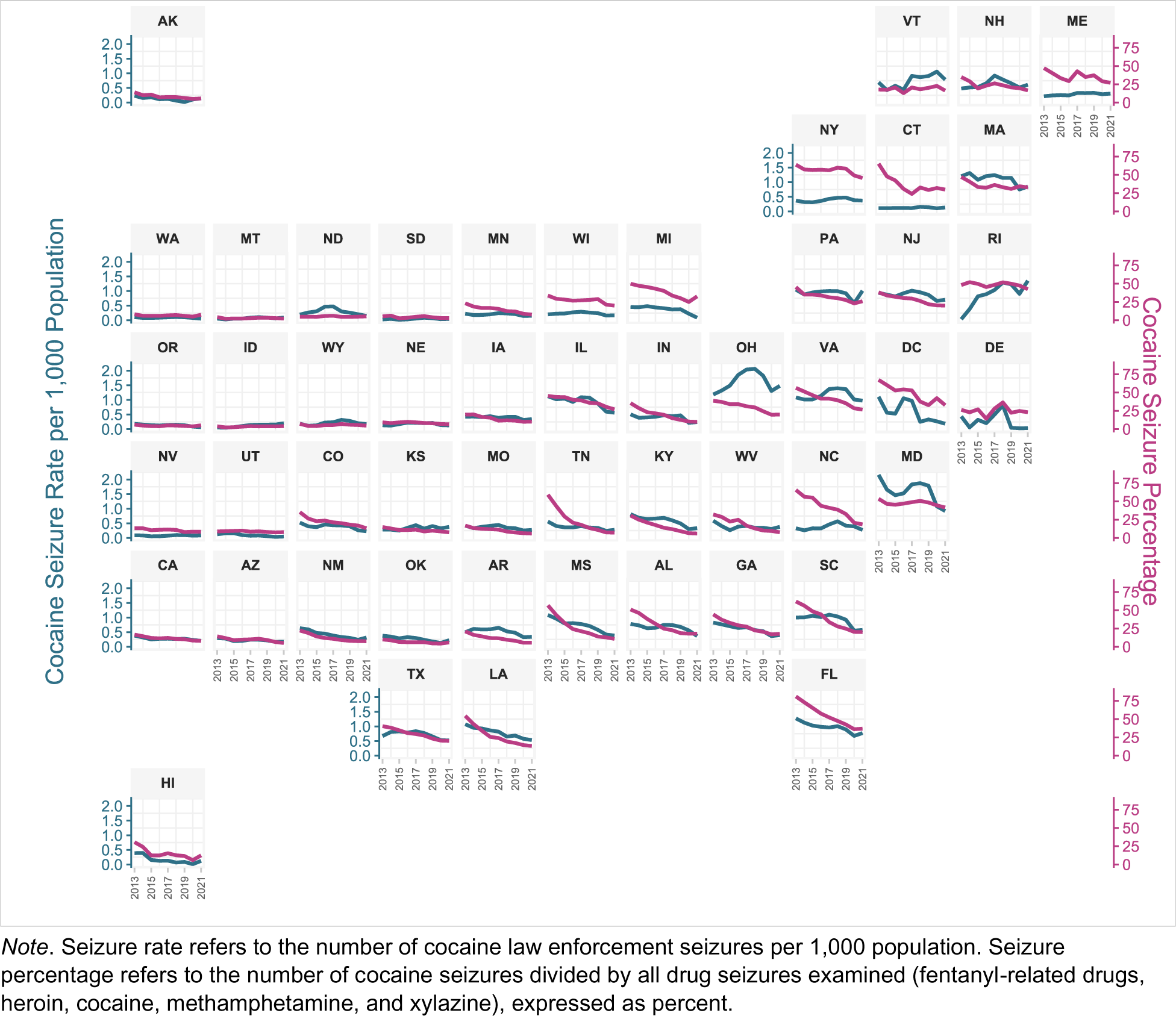
Distributions of Cocaine Seizure Rates/Percentages by State, US, 2013-2021.

**Figure S5.**
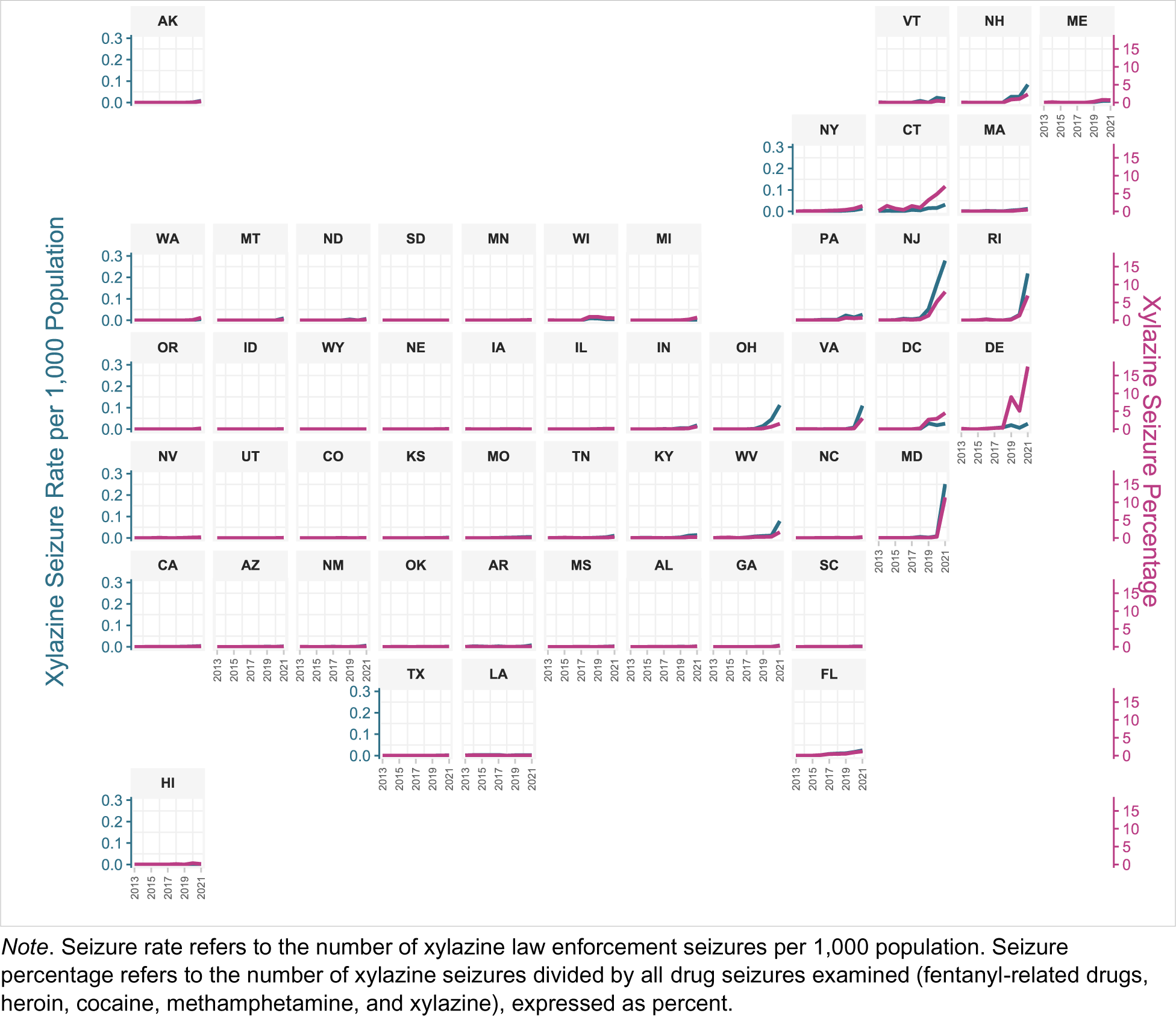
Distributions of Xylazine Seizure Rates/Percentages by State, US, 2013-2021.

**Figure S6.**
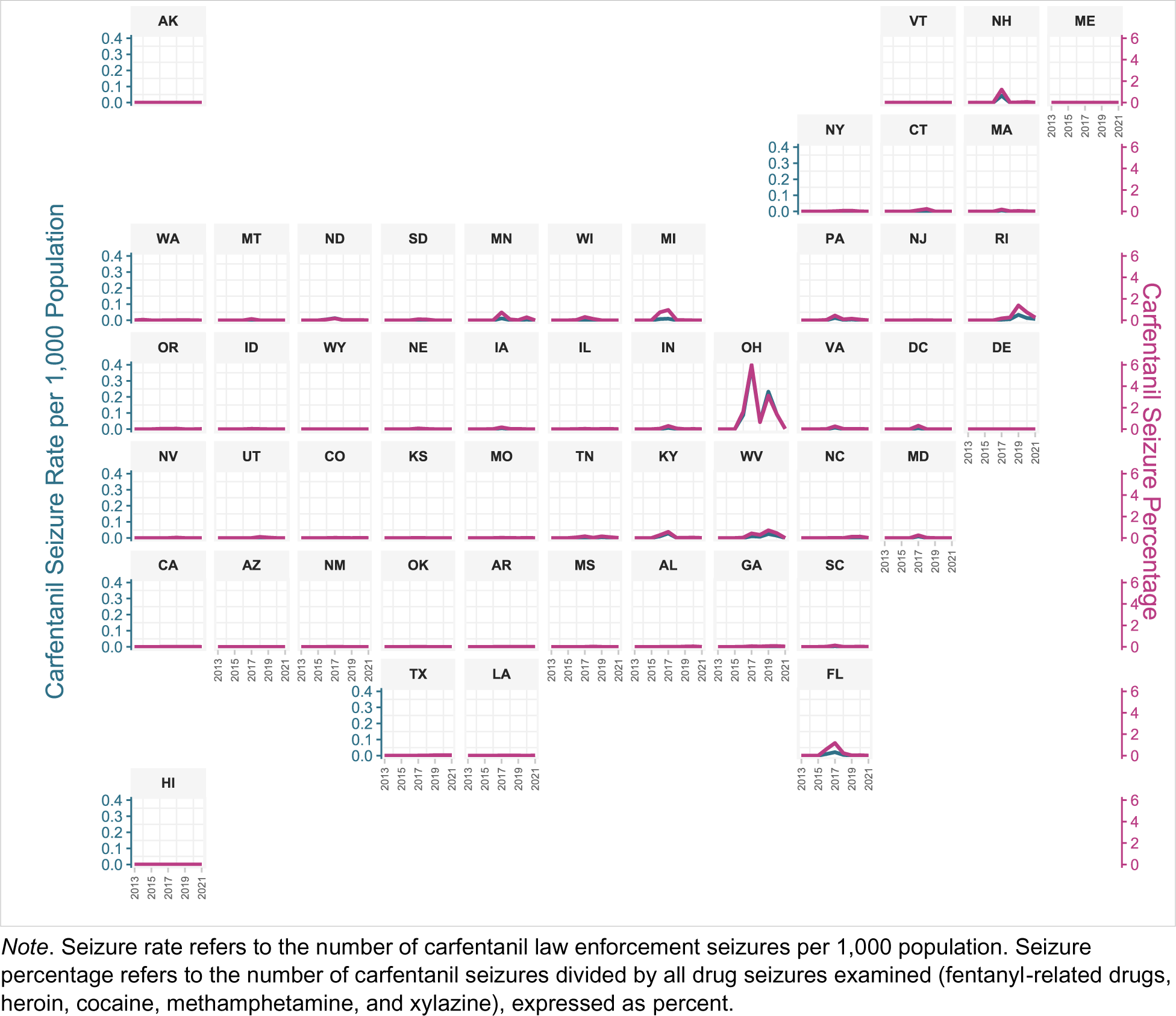
Distributions of Carfentanil Seizure Rates/Percentages by State, US, 2013-2021.

